# The Effect of Sensory Therapy on Upper Extremity Functions and Activities of Daily Living in Patients with Chronic Stroke: A Randomized Controlled Trial

**DOI:** 10.1101/2024.10.02.24314819

**Authors:** Muhammed Rohat Yazıcı, Cigdem Cekmece

## Abstract

**Introduction:** The aim of this study was to investigate the effects of intensive sensory therapy on upper extremity functions, daily activity and life quality of stroke survivors.

**Methods:** This randomized controlled trial included 30 participants aged between 18 and 80 years, who were divided into a sensory-training group (n = 15) and a control group (n = 15). Both groups received 15 sessions of 30 minutes of physical therapy, occupational therapy, and 20 minutes of activity daily living training for three weeks. The ST group also received intensified sensory therapy during each session. Thumb localization, finger shift, and stereognosis tests were performed in both groups only before the start of intervention. The Jebsen Taylor Hand Function Test, Canadian Occupational Performance Measure, Modified Frenchay Scale, and Stroke Specific Quality of Life Scale (SS-QOL) were administered before and after intervention.

**Results:** Statistically significant differences were observed between the ST group and the control group across all tests, except for the Language, Thinking, and Vision parameters of the SS-QOL.

**Conclusion:** These results suggest that sensory therapies applied with conventional interventions increased upper extremity functions, activity daily living performance, participation and quality of life of the stroke survivors.

**Registration:** URL: https://clinicaltrials.gov/study/NCTxxxxxx Unique identifier: NCTxxxxx.

## Introduction

Internationally, stroke remains a leading cause of disability, imposing significant economic burdens on post-stroke care worldwide (Rajsic et al., 2019). A significant portion of stroke survivors, approximately 35%, experience complete or partial dependence in activities of daily living (ADL) (Carey et al., 2016). Stroke survivors commonly face motor, sensory, and cognitive challenges (Hoffmann et al., 2010). These dysfunctions, occurring in chronic stroke, affect both upper and lower extremities, leading to dependence in personal care, work productivity, leisure activities, and social participation, while also impacting personal, family, and work roles (Gillen, 2015). Otherwise the high prevalence and significance of somatosensory function in daily activities, there is limited empirical evidence linking impaired somatosensorial with participation in these activities (Carey et al., 2016).

Post-stroke sensory rehabilitation generally follows two key approaches: explicit sensory- focused training, where patients actively engage in detecting and recognizing sensory stimuli, and implicit sensory-focused training, which involves repeated exposure to sensory stimuli without direct attention or verbal interaction (Doyle et al., 2010; Serrada et al., 2019).

Sensory inputs are fundamental to motor recovery after stroke, as motor movements require the integration of sensory information (Chen et al., 2018). Thus, motor rehabilitation should include sensory training (Doyle et al., 2014a; Doyle et al., 2014b). However, therapies often prioritize motor functions, overlooking the auxiliary role of sensory inputs (Zandvliet et al., 2020). A promising approach is a rehabilitation process that integrates sensory therapy, involving somatosensory, visual, auditory, tactile, vestibular, or multisensory stimulation, alongside targeted movements (Hildebrand et al., 2023). Researchers suggest that sensory therapy, such as peripheral nerve stimulation and muscle tendon vibration, may improve motor performance by increasing corticospinal excitability and expanding the representation of the excited body part (Edwards et al., 2014). Furthermore, restoring sensorimotor interactions in the damaged motor system is beneficial for motor recovery (Laible et al., 2012).

Sensory disruptions in the upper extremity after stroke often lead to persistent challenges in everyday tasks like self-care, household chores, and recreational activities. However, sensory disorders are often overlooked in stroke rehabilitation, with more focus placed on motor function and exercises for the upper and lower extremities (Byrne et al., 2023). Since re-establishing sensory processing and sensorimotor interactions is crucial for motor recovery, sensory-based therapies should be integrated with conventional intervention methods (Aguia- Rojas et al., 2023).

Research on sensory rehabilitation has often focused on either active sensory exercise, such as identifying different tactile stimuli or passive methods like electrical stimulation, hot or cold applications, and pneumatic compression (Walker et al., 2022). Conversely increasing evidence supporting the benefits of specific sensory training, its application in clinical practice remains limited (Doyle et al., 2013a; Doyle et al., 2014b). Therefore, there is a need for more robust evidence supporting the use of sensory therapies in stroke intervention. A deeper understanding of how sensory impairments affect daily life will enable clinicians to design better, individualized rehabilitation interventions. A recent systematic review identified a significant need for standardized sensory retraining protocols tested through rigorous methodologies to ensure their effectiveness in clinical settings (Gavin et al., 2022). Future studies should focus on sensory-based occupational therapy interventions for stroke survivors (Carey et al., 2018; Doyle et al., 2013b). There are theoretical, clinical, and academic gaps in sensory assessment and interventions for individuals with chronic stroke. Accordingly, this study was planned. This randomized controlled study aimed to investigate the effects of sensory-based occupational therapy on upper extremity sensory functions, ADL, and quality of life in chronic stroke patients. Our hypothesis was that sensory therapy would improve upper extremity functions and ADL in stroke survivors.

## Methods

### Study design

This clinical trial followed Consolidated Standards of Reporting Trials (CONSORT) guidelines. It was randomized, single-blind, and conducted in parallel design. This study has been registered with the Clinical Trials Registry (NCTxxxxxxxx). This study conducted at the Kocaeli University Hospital Department of Physical and Rehabilitation Medicine in in Kocaeli, Turkey. The study utilized standardized clinical outcome measures for upper extremity sensory functions, quality of life, and ADL functions in chronic stroke patients. This study was approved by the University Ethical Committee (K). The study CONSORT flow chart is shown in Figure 1.

**Figure 1.**
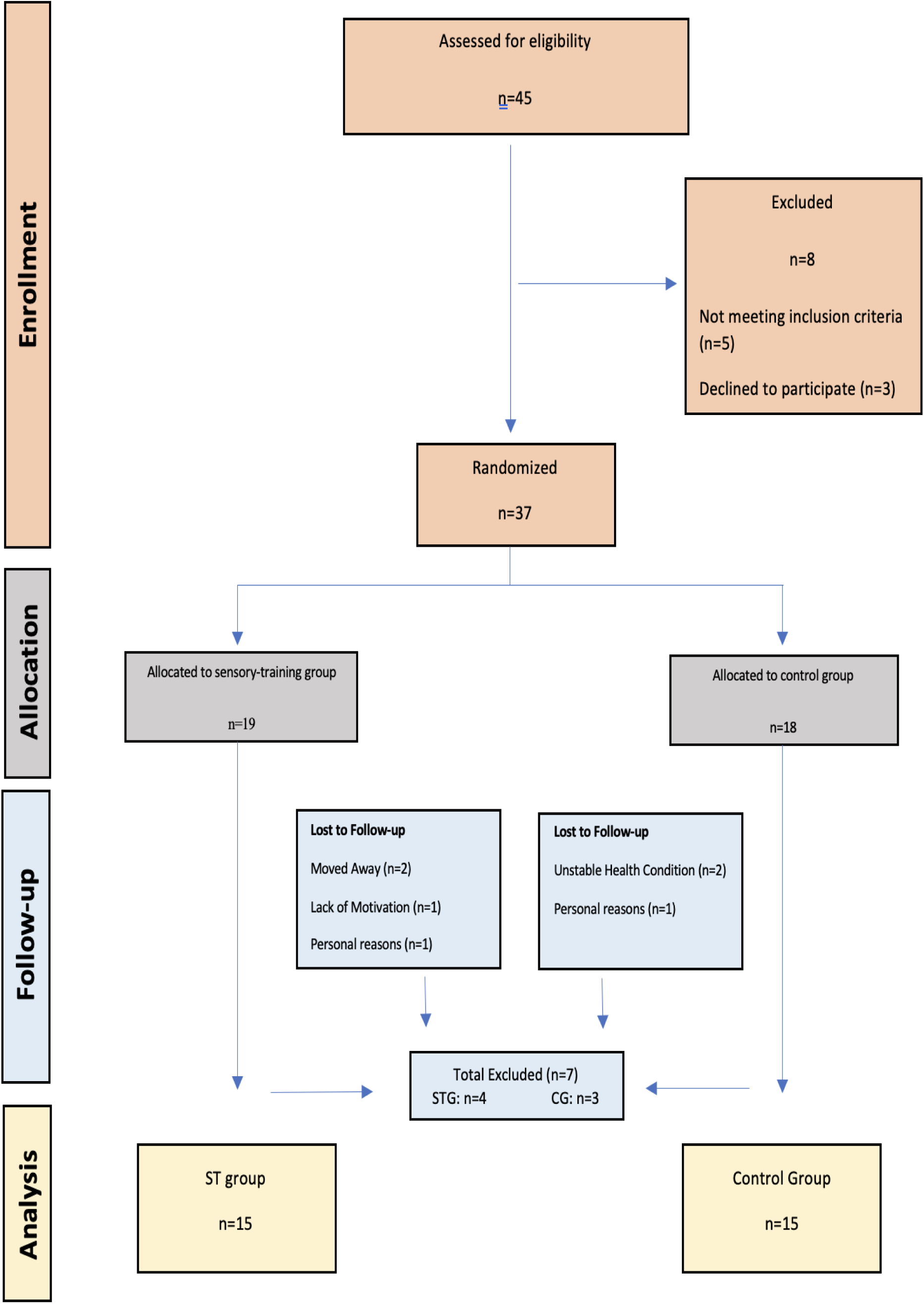
Consort flow diagram of the study.

### Participants

The inclusion criteria were aged between 18 and 80 years, medically stable, a post-stroke interval of at least six months, and a score of more than 24 on the Mini-Mental State Exam. Stroke survivors were not considered for inclusion in the study if they exhibited severe spasticity, indicated by a Modified Ashworth Scale (MAS) score of 3 or higher in upper extremity muscles, had joint limitations (contractures), congestive heart failure, peripheral arterial disease, severe dementia, language impairments, or experienced highly painful conditions like reflex sympathetic dystrophy.

### Allocation

This study was conducted with stroke survivors who were hospitalized at the Physical and Rehabilitation Department of Kocaeli University Hospital between May and September 2022. Participants who completed the assessment were then randomly assigned to intervention groups. Assessments and interventions were conducted in the occupational therapy department of this hospital. Informed written consent was obtained from all participants before participating in the study.

### Randomization and blinding

Simple randomization method was used for the study. The participants were randomly assigned in a 1:1 ratio to the ST group or the control group by a researcher who did not participate in the intervention or the evaluation process. To ensure standardization, all evaluations were made by the same researcher who was blind to the intervention group. Since this researcher is not part of the standard intervention team, she is not aware of this specific intervention modality the patient received, which ensures blinding. Physiotherapists who applied conventional intervention, and occupational therapists who applied occupational therapy, ADL training, and sensory interventions were not blinded to the study.

For randomization, A person who was not part of the study made the allocation. To ensure allocation concealment, our study used an independent, centralized allocation protocol that did not involve any individuals associated with the study. With this method, randomization lists were created and maintained securely in remote locations (in the hospital’s main building), thus reducing the chance of monitoring intervention assignments and the process. With a centralized intervention assignment, a participant who was eligible and consented to the study was assigned to the intervention group (sensory group or control group) by calling the randomization centre to receive the intervention assignment, after recording demographic information and initial assessments. It is clear from this example that neither patients nor researchers can predict the next allocation; This is designed to hide the allocation.

### Intervention

All participants in the study received a 3-week intervention consisting of a total of 15 sessions. While one session of participants in the control group lasted approximately 1 hour, one session of participants in the sensory training group was approximately one and a half hours. The interventions were conducted in person and on an individual basis. The specific content of the interventions in the two groups is outlined below:

All participants received physical therapy, which included hot-pack application, stretching exercises, strengthening exercises, balance, and ambulation training as appropriate. In the occupational therapy program, which was applied to both groups for 30 minutes a day, fine grasping exercises (screwing objects together, removing nails, and placing round rings), rough grip exercises (throwing balls, cup activity, placing big box blocks, and gripping cylinders), bilateral training and teaching activities (hand cycling, climbing ladders, hitting two objects together, reaching for a large ball with both hands) were applied. Following a 5- minute rest period, participants undertook 20 minutes of training in ADL.

The ADL training focused on working on activities identified by participants using the COPM and scales and included activities such as cooking, shaving, taking a shower, walking, and visiting relatives. The aim was that participants would be able to perform their identified activities independently. In addition, ADL training aimed to increase the stroke survivors’ functional independence by making activity modifications and environmental arrangements, such as thickening the fork handle, adding a cup holder, teaching techniques for wearing cardigans, t-shirts, and trousers, and one-handed lacing methods.

In the sensory-training group, the ADL training was followed by a 5-minute rest period, after which participants received sensory therapy for 30 minutes. As sensory therapy, training on recognizing different surfaces, weight transfer, vestibular sensory studies, approximations, deep massage applications, and brushing were performed. In the training for recognizing different surfaces, wooden blocks of surfaces of different textures and hardness were used.

Barbed balls and hard and soft sensory brushes were used for brushing. The stroke survivors’ affected hand and arm were brushed for a duration of 5 minutes. Touching activities, using hot and cold tubes, were also included in the application to create thermal, tactile stimulation in the stroke survivors. After brushing, weight-bearing was performed with the stroke survivors’ arm on the table (1 minute), extended in the sitting position (1 minute), and with the arm leaning against the wall (1 minute) for a total of 3 minutes. Deep massage applications to the affected hand and arm and therapist-assisted approximations were also performed as part of the intervention. Intervention was completed with vestibular sensory studies.

### Assessments and outcomes

Demographic information of the stroke survivors, including age, gender, dominant hand, plegic side, and time after stroke, was recorded. The stroke survivors’ sensory evaluation (Thumb Localization Test, Finger Shift Test, and Stereognosis Test) results were recorded before the intervention. At the beginning and end of the intervention, the Jebsen Taylor Hand Function Test (JTHFT) (Davis Sears & Chung, 2010), Canadian Occupational Performance Measurement (COPM) (Carswell et al., 2004), Modified Frenchay Scale (MFS) (Laclergue et al., 2023), and Stroke-Specific Quality of Life Scale (SS-QOL) (Hakverdioğlu Yönt & Khorshid, 2012), were applied.

### Primary outcomes

Primary outcomes are JTHFT and COPM. JTHFT consists of seven subsections (writing, turning cards, moving small objects, simulated feeding, stacking checkers, moving empty cans, and moving full cans). As some of the stroke survivors participating in the study were illiterate, the first item of JTHFT, the act of “writing”, was excluded from the evaluation.

COPM is a client-cantered, and semi structured assessment tool used to identify and evaluate the stroke survivors perceived performance and satisfaction in everyday activities. Patients are asked about the activities in which they have problems in these areas, their performance and satisfaction levels. Performance and importance levels are determined by the patient on a 1-10 scale as “Not at all important” (1) and “Very important” (10).

### Secondary outcomes

Secondary outcomes are MFS and SS-QOL. MFS is a scale that evaluates stroke survivors’ upper extremity functions and instrumental ADL (drawing a straight line with a ruler, opening a jar compartment, attaching a peg, combing hair, etc.). Scoring for each activity was done as 0 (no movement), 5 (completed the task) and 10 (normal movement).

SS-QOL consists of 12 sub-sections including energy, family roles, language, mobility, mood, personality, self-care, social roles, thinking, upper extremity function, vision and work/productivity and a total of 49 questions; It is graded with Likert type scoring ranging from 1 to 5.

### Sample size calculation

The sample size for the trial was calculated based on the Jebsen Taylor Hand Function Test (JTHFT) as the primary indicator, with an anticipated mean effect size of d = 1.42, as observed in previous studies (Allgöwer & Hermsdörfer, 2017; Krumlinde-Sundholm et al., 2019). A sample size of 15 patients in each group was determined using G*power Software (version 3.1.9.6) with an alpha level (α) of 0.05 and power (1–β) of 0.950. Accordingly, a total of 30 participants were included in the study and control groups.

### Statistical Analysis

Statistical analysis was done with IBM SPSS, version 20.0 (IBM Corp, Armonk, NY, USA). Normality of data set distribution was evaluated with the Shapiro-Wilk Test. Normally distributed numerical variables are given as mean±standard deviation, non-normally distributed numerical variables as median (25th-75th percentile), and categorical variables as frequency (percentage). The difference between the groups was determined by independent samples t-test and Mann-Whitney U test, as appropriate. Differences between dependent samples were analysed by paired t-test and Wilcoxon signed-rank test. A p<0.05 was considered sufficient to indicate statistical significance when testing two-sided hypotheses.

### Results Enrollment flow

Of the 45 stroke patients assessed, five were excluded due to not meeting the inclusion criteria and three refused to participate. The remaining 37 were randomised to study (n = 19) or control (n = 18) groups. Two participants moved away, one due to lack of motivation, two due to personal reasons, and two due to unstable health were lost to follow-up. The final analysis included 15 participants in the sensory training group and 15 in the control group, as shown in Figure 1, which illustrates the flow of participants.

### Pre-intervention characteristics

Baseline characteristics were comparable between the ST and control groups. (p > 0.05). The demographic information of the stroke survivors regarding gender, dominant hand, hemiplegic side, and time after stroke is given in Table 1. Sensory evaluations of the stroke survivors were examined before the intervention, both groups were similar in terms of all sensory functions (Table 1) (p>0.05). The outcome of JTHFT, COPM, MFS, and SS-QOL of stroke survivors before intervention are given in Table 2. No significant differences were found between the two groups in the pre-intervention evaluations (p>0.05).

**Table 1.**
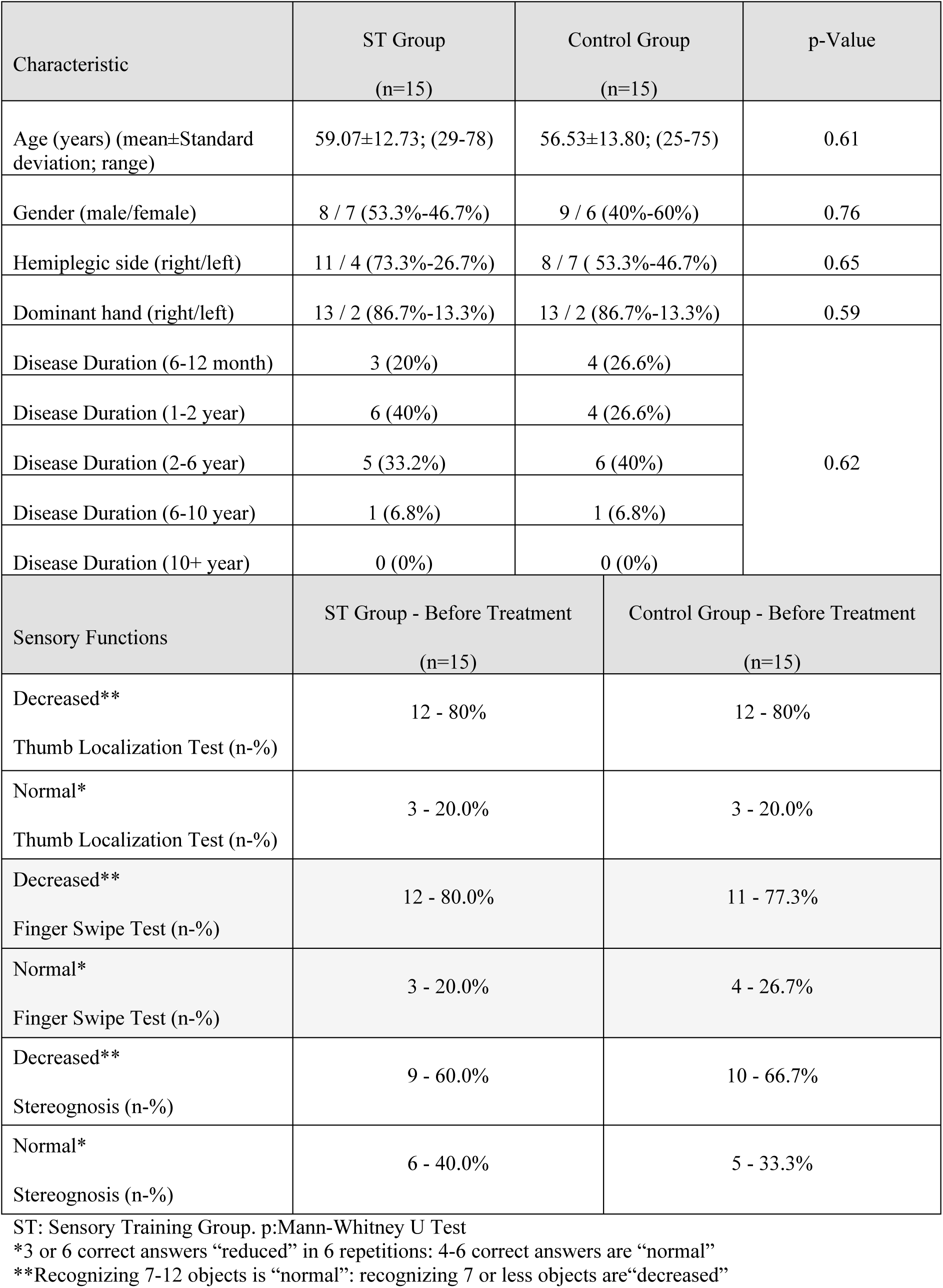
Characteristic and sensory functions of study participants (mean, SD, range, and number)

**Table 2.** Clinical outcome measures at before treatment (BT), and after treatment (AT) in participants.

| Outcome | Sensory training group |  | Control group |  | Intragroup comparison |  | Intergroup comparison |  |
| --- | --- | --- | --- | --- | --- | --- | --- | --- |
|  | BT | AT | BT | AT | ST group (p<sup>a</sup>) | Control group (p<sup>b</sup>) | BT (p<sup>c</sup>) | AT (p<sup>d</sup>) |
| COPM Performance | 3.2 (2.8-3.6) | 5.6 (5 - 6.8) | 3.8 (3.2-4) | 4 (4 - 5) | <0.01 | <0.01 | 0.18 | <0.01 |
| COPM Satisfaction | 4 (3.2-4.6) | 6.2 (5.6 - 7) | 4 (3-5) | 5 (4.2 - 6) | <0.01 | <0.01 | 0.80 | <0.01 |
| COPM Total | 7.2 (6.6 - 7.8) | 12.2 (10.6 - 13) | 7.6 (6.4 - 9) | 9.8 (8.2 - 11) | <0.01 | <0.01 | 0.57 | <0.01 |
| JTHFT/ Turning cards | 40.14 (18.53-52.65) | 28.01 (10.36-34.73) | 38.87 (25.17-49.1) | 37.43 (24.42-47.9) | <0.01 | <0.01 | 0.87 | 0.02 |
| JTHFT/ Moving small objects | 47.5 (22.87-59.49) | 34.24 (12.97-39.6) | 49.53 (38.2-57.49) | 48.7 (35.96-55.03) | <0.01 | <0.01 | 0.71 | <0.01 |
| JTHFT/ Simulated feeding | 40.65 (21.66-50.66) | 30.12 (14.27-39.62) | 40.55 (33.83-48) | 38.67 (31.45-47.03) | <0.01 | <0.01 | 0.90 | 0.01 |
| JTHFT/ Stacking checkers | 29.46 (20.09-48.49) | 25.3 (16.56-33.46) | 38.84 (25.88-53.99) | 36.12 (23.56-49.8) | <0.01 | <0.01 | 0.32 | 0.04 |
| JTHFT/ Moving empty cans | 29.56 (17.82-40.6) | 23.18 (11.88-27.28) | 36.22 (27.56-43.51) | 35.74 (26.59-40.17) | <0.01 | <0.01 | 0.25 | <0.01 |
| JTHFT/ Moving full cans | 33.9 (20.07-39.72) | 25.37 (12.72-32.03) | 39.59 (31.9-54.12) | 37.59 (28.3-50.54) | <0.01 | <0.01 | 0.13 | <0.01 |
| MFS1/ Opening jar lid | 3 (3-7) | 9 (8-10) | 7 (3-7) | 6 (4-8) | <0.01 | <0.01 | 0.10 | <0.01 |
| MFS2/ Drawing lines with a ruler | 5 (4-6) | 8 (5-9) | 5 (4-6) | 4 (3-7) | <0.01 | 0.65 | 0.35 | <0.01 |
| MFS3/ Big cup holding | 6 (3-8) | 8 (8-10) | 6 (3-8) | 5 (4-8) | <0.01 | <0.01 | 0.16 | 0.02 |
| MFS4/ Small cup holding | 5 (2-7) | 7 (6-9) | 5 (2-7) | 5 (3-7) | <0.01 | <0.01 | 0.23 | 0.01 |
| MFS5/ Drinking water | 4 (3-5) | 7 (5-9) | 4 (3-5) | 5 (2-7) | <0.01 | 0.24 | 0.79 | 0.01 |
| MFS6/ 3 pegs install | 4 (2-5) | 6 (4-8) | 4 (2-5) | 2 (2-5) | <0.01 | 0.19 | 0.98 | <0.01 |
| MFS7/ Hair combing | 4 (3-7) | 5 (4-9) | 3 (2-5) | 4 (3-7) | <0.01 | <0.01 | 0.08 | 0.02 |
| MFS8/ Squeezing toothpaste | 5 (4-7) | 8 (5-9) | 3 (3-5) | 4 (4-6) | <0.01 | <0.01 | 0.10 | 0.01 |
| MFS9/ Using a fork and knife | 4 (3-7) | 7 (5-9) | 3 (2-6) | 5 (3-7) | <0.01 | <0.01 | 0.51 | 0.02 |
| MFS10/ Using a broom | 5 (2-6) | 8 (6-9) | 4 (2-6) | 5 (3-7) | <0.01 | <0.01 | 0.96 | 0.03 |
| SSQLS/ Energy | 6 (4-7) | 10 (8-14) | 5 (3-8) | 7 (5-10) | <0.01 | 0.01 | 0.51 | 0.02 |
| SSQLS/ Family roles | 5 (5-8) | 8 (7-11) | 5 (4-5) | 7 (5-8) | <0.01 | <0.01 | 0.14 | 0.02 |
| SSQLS/ Language | 15 (10-25) | 20 (13-25) | 15 (6-25) | 19 (10-25) | <0.01 | 0.01 | 0.93 | 0.74 |
| SSQLS/ Mobility | 12 (9-16) | 23 (19-26) | 14 (9-23) | 15 (11-23) | <0.01 | 0.08 | 0.22 | 0.02 |
| SSQLS/ Mood | 15 (10-19) | 20 (18-24) | 12 (10-18) | 15 (13-20) | <0.01 | <0.01 | 0.53 | <0.01 |
| SSQLS/ Personality | 7 (4-12) | 11 (8-15) | 6 (3-12) | 8 (6-13) | **<0.01** | **<0.01** | 0.41 | **0.03** |
| SSQLS/ Self-care | 15 (11-17) | 20 (18-24) | 14 (11-17) | 16 (13-17) | **<0.01** | **<0.01** | 0.52 | **<0.01** |
| SSQLS/ Social roles | 9 (7-9) | 13 (12-15) | 7 (5-9) | 10 (8-13) | **<0.01** | **<0.01** | 0.07 | **<0.01** |
| SSQLS/ Thinking | 6 (4-9) | 10 (8-15) | 6 (3-13) | 10 (5-14) | **<0.01** | **<0.01** | 0.75 | 0.16 |
| SSQLS/ Upper Extremity Function | 12 (7-15) | 19 (13-23) | 11 (9-14) | 14 (10-16) | **<0.01** | **<0.01** | 0.96 | **0.01** |
| SSQLS/ Seeing | 11 (9-15) | 13 (11-15) | 15 (12-15) | 15 (12-15) | 0.08 | 0.06 | 0.28 | 0.19 |
| SSQLS/ Work-productivity | 5 (3-6) | 8 (6-11) | 5 (4-6) | 6 (4-8) | **<0.01** | **0.03** | 0.93 | **0.03** |
| SSQLS/ Total | 115 (104-159) | 174 (163-216) | 118 (97-141) | 148 (120-154) | **<0.01** | **<0.01** | 0.77 | **<0.01** |

Data were presented as median (p25,p75). Bold indicate values that are statistically significant. ST= Sensory Training BT= Before Treatment AT= After Treatment
COPM= Canadian Occupational Performance Measure JTHFT= The Jebsen Taylor Hand Function Test MFS= Modified Frenchay Scale SS-QOL= Stroke Specific Quality of Life Scale
p<sup>a</sup> = value for between groups comparisons (Mann-Whitney U test)
p<sup>b</sup> = value for between groups comparisons (Mann-Whitney U test)
p<sup>c</sup> = value for within groups comparisons (Wilcoxon signed rank test)
p<sup>d</sup> = value for within groups comparisons (Wilcoxon signed rank test)

### Changes from post-intervention

The assessments of JTHFT, MFS, COPM and SS-QOL of stroke survivors after intervention are given in Table 2. Scores in the post-intervention JTHFT categories were significantly different between both groups: turning cards (p=0.026), moving small objects (p=0.017), simulated feeding (p=0.017), stacking checkers (p=0.044), moving empty cans (p=0.003) and moving full cans (p=0.007) (Table 2). Moreover, the differences of all JTHFT parameters before and after intervention were taken (intervention response), statistical analysis was made and added graphically (Figure 2). Significant differences in improvement were also found in the ST group for COPM performance (p=0.001) and satisfaction (p=0.006) values when compared to the control group (Table 2). Furthermore, according to the COPM assessment, the activities that participants in both the ST group and the control group had problems with are given in Table 3.

**Figure 2.**
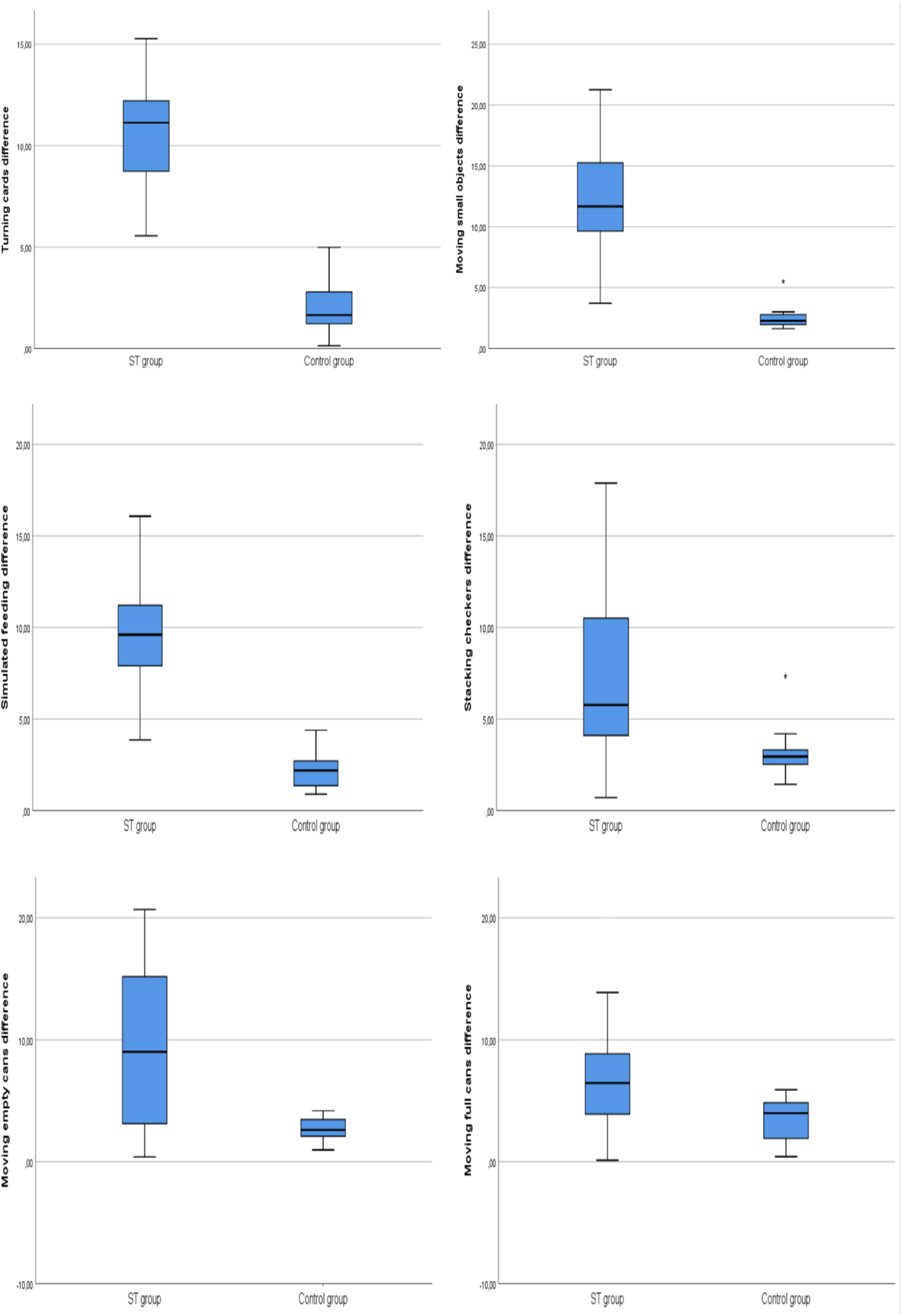
Analysis graph of pre/post treatment differences of JTHFT parameters.

**Table 3.** Activities in which participants have problems according to COPM.

| Activities |  | ST Group<br>(n=15) |  | Control Group<br>(n=15) |  |
| --- | --- | --- | --- | --- | --- |
|  |  | n | % | n | % |
| Self-Care Activities | Brushing teeth | 3 | 20 | 4 | 26.6 |
|  | Shaving | 4 | 26.6 | 1 | 6.6 |
|  | Take a shower | 7 | 46.6 | 9 | 60 |
|  | Cut nail | 4 | 26.6 | 2 | 13.3 |
|  | Dressing-Undressing | 5 | 33.3 | 4 | 26.6 |
| Productivity/<br>Work-Related Activities | Cooking | 5 | 33.3 | 3 | 20 |
|  | Go to work | 3 | 20 | 4 | 26.6 |
|  | Cleaning up | 4 | 26.6 | 5 | 33.3 |
| Leisure and Social Participation Activities | Take a walk | 5 | 33.3 | 3 | 20 |
|  | Spending time with grandchildren | 3 | 20 | 4 | 26.6 |
|  | Spending time with family | 3 | 20 | 4 | 26.6 |
|  | Knit | 2 | 13.3 | 1 | 6.6 |
|  | Visiting relatives/friends | 3 | 20 | 4 | 26.6 |

All the MFS scores of the ST group and control groups were compared after the intervention, there was a significant difference in the activities of all parameters after the intervention (p<0.05) (Table 2). When the SS-QOL results of the ST group and the control group were compared after intervention, a statistically significant difference was found in favour of the ST group (p<0.05), except for language, thinking and visual domains (p>0.05) (Table 2).

## Discussion

This randomized controlled study explored the impact of sensory therapy on upper extremity functions, ADL performance, and quality of life in stroke survivors. After a three-week intervention, the sensory therapy group showed significant improvements in motor function and quality of life compared to the control group, who received conventional therapy without sensory training. Additionally, the sensory therapy group demonstrated enhanced ADL performance and satisfaction. These findings highlight the potential of sensory therapy as a complementary approach to traditional stroke rehabilitation.

JTHFT was utilized to assess motor function in the plegic upper extremity. This test includes tasks involving common grips used in daily activities and evaluates the speed with which individuals perform these tasks. Unlike self-reported scales, JTHFT provides objective assessment results that are independent of a participant’s perception of performance. Good performance in this test requires the accurate integration of complex sensorimotor information (Duff et al., 2006). In everyday activities, people frequently encounter tasks requiring grip, reach, and shoulder stabilization. For stroke survivors, proprioceptive sensory impairments often impede the coordinated execution of these tasks. The greater improvements seen in these activities in the sensory therapy group strengthen the argument that stroke survivors receiving this additional therapy may use their upper extremity more functionally in ADL. This aligns with earlier studies that found positive outcomes when sensory rehabilitation was added to conventional interventions for upper extremity motor function (Allgöwer & Hermsdörfer, 2017; Blennerhassett et al., 2007). In this study, a statistically significant improvement in motor function was found in the sensory therapy group, particularly in tasks involving recognition of different textures.

Bernard-Espina et al. (Bernard-Espina et al., 2021) also demonstrated the importance of multisensory integration, including hand motor control, vision, and proprioception, in motor function recovery following stroke. They concluded that stroke affects both motor and sensory functions. In a randomized controlled trial conducted by Allgöwer and Hermsdörfer, JTHFT was used similarly to our study, and the researchers found that fine motor skills for grasping, holding, and manipulating objects require interaction among multiple sensorimotor systems (Allgöwer & Hermsdörfer, 2017).

A semi-structured interview using the COPM was conducted to assess stroke survivors’ activity performance, satisfaction, and participation. Both groups showed significant improvement in all COPM subfields, but post-intervention comparisons revealed a significant advantage for the sensory therapy group. Hejazi-Shirmard et al., in their comparison of top-down and bottom-up rehabilitation, found COPM scores like those in our study (Hejazi-Shirmard et al., 2024). According to COPM results, an improvement of 2 points or more is considered moderate to high clinical improvement, which correlates with significant performance changes in stroke survivors (Law et al., 1994). In our study, the sensory therapy group showed increases of 2.34 points in performance and 2.45 points in satisfaction after intervention, while the control group showed increases of only 0.7 and 1.2 points, respectively. This level of improvement (>2.0) suggests that combining sensory therapy with patient-centred ADL education effectively enhances ADL performance in stroke survivors. Common areas of limitation identified in stroke survivors included self-care (e.g., brushing teeth, showering), productivity-work (e.g., cooking, cleaning), and leisure (e.g., walking). COPM is notable for its ability to assess participation levels, and in our study, stroke survivors expressed a desire to spend more time with family and engage in these activities as part of their intervention.

While changes in various domains impact survivors’ performance in daily life, there are limited studies on the effectiveness of sensory-based rehabilitation practices for ADL improvement (Alwawi et al., 2024). We utilized the MFS to evaluate instrumental ADLs.

MFS showed significant improvements in the sensory therapy group. However, 8 of 15 stroke survivors in the sensory therapy group and 4 of 15 in the control group achieved the expected level of performance, while 2 in the sensory therapy group and 1 in the control group exceeded their goals. Studies by Eghlidi et al. and Derakhshanfar et al. have similarly demonstrated significant improvements in ADL performance through sensory-based rehabilitation added to conventional interventions (Derakhshanfar et al., 2021; Eghlidi et al., 2015).

Following a stroke, many survivors experience functional loss, including balance and walking difficulties, upper extremity dysfunction, speech disorders, swallowing problems, and sensory loss, which negatively affect daily activities and quality of life (Ghrouz et al., 2024). Quality of life in stroke survivors is influenced by physical, functional, cognitive, and social factors (Rocha et al., 2021). In this study, we used the SS-QOL scale to assess the impact of sensory therapy. Post-intervention comparisons showed significant improvements in the sensory therapy group across all domains except “Language,” “Thinking,” and “Sight.” Although there are studies examining the effectiveness of sensory therapy in stroke survivors, there is no study evaluating the effect of sensory therapy on the quality of life of stroke survivors. The results of the present study showed that sensory training has positive effects on upper extremity functionality and independence, participation in ADL, and thus improved quality of life in stroke survivors.

Our findings suggest that sensory training positively influences upper extremity function, ADL participation, and quality of life. Despite the common emphasis on motor function recovery in early stroke rehabilitation, sensory function interventions are often neglected. Our study, along with others, suggests that sensory training should be incorporated into early rehabilitation protocols.

### Limitation

One limitation of our study is the short follow-up period of three weeks, which leaves the long-term effectiveness of the intervention unexamined. Another limitation is the relatively small sample size, which could be addressed in future research with larger, prospective studies that include long-term follow-up. Additionally, while both groups showed improvement, the greater effect observed in the sensory therapy group complicates the identification of which specific sensory stimuli contributed to the outcomes. Future studies should consider isolating individual sensory stimuli to better understand their distinct contributions.

## Conclusions

This randomized controlled study suggests that thorough assessment of sensory functions and the integration of sensory therapy into routine interventions are crucial for enhancing functionality in stroke survivors. Moreover, participants who received sensory therapy reported significant improvements in quality of life. We believe that the variety of materials, devices, and environments used in sensory therapy plays a key role in promoting recovery in stroke survivors.

## Key findings

- This study will provide a resource and encourage occupational therapists working in stroke rehabilitation to include sensory processes in their therapies.
- This study may create awareness among occupational therapy practitioners about the importance of sensory therapy in stroke rehabilitation.
- This study may encourage occupational therapists to work with stroke survivors for sensory therapy and allow planning interventions and comparing results.

## Declarations

The authors confirm that there is no conflict of interest.

## Availability of data and material

The datasets created and/or analyzed during the current study are not publicly available due to [participants’ deprivation of personal information] but are available from the corresponding author upon reasonable request.

## Ethics approval

The study was approved by the local institutional ethical board (Ethics Committee at Kocaeli University) and it was examined by the ministry of health and was found in accordance with the regulation (Number: (KAEK 2021/04.39).).

All the procedures performed were in accordance with the ethical standards of the national research standards and with the 1964 Helsinki declaration and its later amendments. Informed consent was obtained from all the individual participants included in the study.

## Statement of contributorship

All authors contributed to the development of the study methodology, data collection, and analysis. All authors participated in writing, reviewing, and editing the manuscript, and approved the final version.

## Data availability statement

Data are available upon reasonable request from the corresponding author and are otherwise restricted for ethical considerations, as it was instructed by the ethical committee.

## Acknowledgements

The authors would like to thank all the individuals who participated in this study.

## Consent to participate

All respondents signed online informed consent forms for participation.

## Contributorship

Conceptualization, Ç.Ç, and M.R.Y; methodology, Ç.Ç.; software, Ç.Ç, and M.R.Y.; formal analysis, Ç.Ç, and M.R.Y.; investigation, M.R.Y.; resources, M.R.Y.; data curation, Ç.Ç, and M.R.Y.; writing—original draft preparation, Ç.Ç.; writing—review and editing, Ç.Ç; visualization, Ç.Ç. All authors have read and agreed to the published version of the manuscript.

